# Feasibility and acceptability of Narrative Exposure Therapy to treat individuals with PTSD who are homeless or vulnerably housed: A pilot randomized controlled trial

**DOI:** 10.1101/2021.11.08.21266074

**Authors:** Nicole E. Edgar, Alexandria Bennett, Nicole Santos Dunn, Sarah E. MacLean, Simon Hatcher

## Abstract

**Background:** Annually, there are least 235,000 individuals experiencing homelessness in Canada. These individuals are more likely to have complex health issues, including mental health issues such as post-traumatic stress disorder (PTSD). Diagnosed PTSD rates in the homeless are more than double that of the general population, ranging between 21% and 53%. In the homeless population, complex PTSD (cPTSD) appears to be more common than PTSD. One treatment option for cPTSD is Narrative Exposure Therapy (NET), a brief trauma focused psychotherapy which attempts to place the trauma within a narrative of the person’s life. Previous studies suggest NET may be an effective option for those who are homeless. In this study, our primary aim was to assess the feasibility and acceptability of delivering community-based NET to individuals with PTSD who were homeless or vulnerably housed.

**Methods:** This pilot randomized controlled trial (RCT) enrolled participants who were 18 years of age or older, currently homeless or vulnerably housed, and with active symptoms of PTSD. Participants were randomized to either NET alone or NET plus the addition of a genealogical assessment. Demographic and clinical data were collected at the baseline visit. Symptoms of PTSD, drug use and housing status were re-assessed at follow-up visits. Rates of referral, consent and retention were also examined as part of feasibility.

**Results:** Twenty-two potential participants were referred to the study. Six were not able to be contacted, one was excluded prior to contact, and the remaining 15 consented to participate. Of these, one was a screen failure and 14 were randomized equally to the treatment arms. One randomized participant was withdrawn for safety. The main point of attrition was prior to starting therapy (3/13). Once therapy was initiated, retention was high with 80% of participants completing all six sessions of therapy. Seven participants completed all follow-up sessions.

**Conclusion:** Delivering NET in a community-based setting and completing genealogical assessments was both feasible and acceptable to those who are homeless or vulnerably housed. Once therapy had been initiated, participants were likely to stay engaged. A large RCT should be conducted to evaluate effectiveness and feasibility on an increased scale.

**Trial Registration:** clinicaltrials.gov, identifier NCT03781297. Registered: December 19, 2018, https://clinicaltrials.gov/ct2/show/NCT03781297.

**Key messages regarding feasibility:**

1. *What uncertainties existed regarding the feasibility?* Narrative Exposure Therapy (NET) is an individual trauma focused psychotherapy recommended in guidelines for the treatment of post-traumatic stress disorder (PTSD). However, in people who are homeless or vulnerably housed, there have been no randomised controlled trials of trauma focused therapies for PTSD. We wanted to find out if it was feasible to recruit and retain people who were homeless with PTSD in a randomized controlled trial of a trauma informed therapy. We wanted to test the acceptability and feasibility of offering NET alone compared to NET plus a genealogical assessment. We also wanted to see if it was acceptable and practical to incorporate a genealogical assessment as part of NET.
2. *What are the key feasibility findings?* The key feasibility finding is that it is feasible to recruit and retain people who are homeless into a randomized controlled trial of a trauma informed therapy. However, recruitment could be improved by a better process for engaging potential participants between referral and enrollment as about a third of the referred population were lost at this stage. Not having trained therapists available also delayed recruitment.
3. *What are the implications of the feasibility findings for the design of the main study?* We will explicitly develop materials for use at the referral step by potential referrers. This will include an online training video which will address issues of trust and how to address them when discussing the potential study. We will also develop a process for training and recruiting therapists. Since this study was completed, we have done two further training workshops to create a pool of potential therapists in Ottawa. We will also engage with another site in Ontario to widen the population base of potential participants.

## Introduction

### Background

Homelessness is a rapidly growing problem in Canada with at least 235,000 individuals experiencing homelessness every year(1,2). In reality this number is likely 3 to 4 times higher to account for individuals with no real prospect of permanent housing options or “hidden homeless”(1). In Canada, point in time counts have repeatedly shown year over year increases in homelessness(2). Exacerbated by a nationwide housing crisis and a global pandemic, accompanied by a reduction in shelter beds and social services, 2021 reports have shown that the situation is continuing to worsen(3–5).

Individuals experiencing homelessness are more likely to have poor overall physical and mental health, increased mortality rates and to experience more barriers to accessing healthcare(6,7) compared to their housed counterparts. In one study nearly 50% of respondents indicated having 3 or more physical health conditions, and 52% indicating having a mental health diagnosis(6,8). Individuals who are homeless are more likely to be exposed to infectious diseases, such as tuberculosis or hepatitis A (6); less likely to seek early treatment and less likely to receive care equivalent to those who are housed (9–12); and face additional challenges with adherence to treatment regimens (13).

### Trauma in the Homeless

Exposure to trauma is a nearly universal experience among the vulnerably housed. It is estimated that as many as 91% of individuals who are homeless have experienced at least one traumatic event(14) and up to 99% have experienced childhood trauma(15,16) Diagnosed post-traumatic stress disorder (PTSD) rates in the homeless are significantly higher than the Canadian population, ranging between 21% and 53%(17–20), compared with a lifetime prevalence of 9.2%(21) in the general population. In addition to trauma before becoming homeless, the experience of being homeless increases the risk of exposure to traumatic events (22). Many who are homeless are exposed to violence, with previous work showing 40% of individuals reporting being assaulted and 21% of women reporting being raped in the previous year (6). Lastly, the experience of being homeless is itself traumatic due to the loss of shelter, safety, stability, and, often, social supports. Thus, being homeless continues to re-traumatize and victimize the individual(23).

In ICD-11, a distinction is made between PTSD and complex PTSD (cPTSD). cPTSD is characterised by experiencing trauma that is prolonged or repetitive from which escape is difficult or impossible (for example, repeated childhood sexual or physical abuse)(24). This results in the symptoms of PTSD plus problems in affect regulation, negative self-beliefs and difficulty sustaining relationships. In the homeless population cPTSD appears to be more common than PTSD, with one survey of 206 homeless adults finding 60% diagnosed with cPTSD and 16% with PTSD(25,26).

This subsequently impacts how individuals engage in health care services, with individuals often experiencing distrust of both people, including healthcare providers, and services(27). It may also lead to self-medication with street drugs to address the symptoms of PTSD. There are also systemic barriers to accessing care which include difficulties finding transportation to appointments, institutional rules that effectively ban people who are homeless, feelings of stigmatization(9,11), having proof of health insurance, and access to no-cost mental health services(28). Most recently, the shift to primarily virtual mental healthcare has further isolated this population from accessing services. Providing therapy in this population should consider the challenges of structural exclusion that this population faces with respect to healthcare services and providers, as well as the unique symptoms associated with cPTSD.

### Narrative Exposure Therapy

Narrative Exposure Therapy (NET) is a brief trauma focused psychotherapy and was developed based on principles derived from exposure therapy, cognitive behaviour therapy and testimony therapy(29). NET attempts to place the trauma within a narrative of the person’s life. This therapy has been evaluated in traumatized populations with a focus on survivors of conflict and organized violence(29). NET is recommended for treatment of PTSD in several guidelines, such as the American Psychological Association guidelines (30) and the National Institute for Health and Care Excellence guidelines(31). There are three therapeutic components which consist of education about the effects of trauma, constructing a biography and narration of traumatic events. The autobiography is recorded by the therapist and is built upon with each subsequent reading. A focus of the therapy is to integrate the generally fragmented reports of traumatic experience into a coherent narrative and to bring about the habituation of emotional responses to reminders of the traumatic event(29). There have been no trials of NET in homeless adults, although one study of NET with 32 street children found a reduction in self-reported offences(32). Anecdotal evidence of using this approach in the homeless population suggests that constructing an autobiography helps to give meaning to problems and provides the initial steps in constructing a core sense of belonging and identity. There is also some evidence that NET may have advantages in treating complex traumatization seen in disadvantaged populations compared to typical first line therapy, such as Prolonged Exposure Therapy(33).

### Incorporation of Genealogy

Genealogy has been used in family therapy(34) and counseling(35) to promote identity(36) and develop connections to ancestors. This can improve relationships with living relatives and potentially address the negative self-beliefs which are part of cPTSD. A systematic review of the acceptability of health and social interventions for people who were homeless found that having a positive self-identity improved links to services(27). The link to those who have gone before is a common theme in indigenous health(37). Previous experience of using problem solving therapy, with a focus on a sense of belonging, in Māori in New Zealand who had presented to hospital with intentional self-harm resulted in improved outcomes after a year compared to usual care. The focus on sense of belonging helped to re-frame individuals’ narrative beyond the immediate family. The metaphor used by participants was that knowing about previous generations helped to deepen their roots so they were less likely to be blown over by life’s storms(38).

### This Trial

As of February 2021, there were no randomized controlled trials of trauma focused therapies in people who are homeless with PTSD. The aim of this study is to test the feasibility and acceptability of delivering community-based NET to individuals with PTSD who were homeless or vulnerably housed. We also extended the option of genealogist support to evaluate the potential impact of this experience on the development of their narrative and self-identity. This trial forms part of the preparation phase of a multi-phase optimization strategy (MOST) for developing and delivering treatment for PTSD in people who are homeless using trauma informed care. Using the outcomes from this study, we will design a full-scale RCT to evaluate and optimize a model of trauma-informed care incorporating NET for the treatment of PTSD in the homeless.

## Methods

### Trial Design

We conducted a single center feasibility, open-label pilot randomized controlled trial with two parallel arms in Ottawa, Canada. The two arms were NET alone (NET) or NET plus a genealogical assessment (NET+G). This trial is reported using the Consolidated Standards of Reporting Trials extension for Feasibility and Pilot Trials (CONSORT)(39).

### Participants

Participants were individuals with PTSD, diagnosed or suspected, who were experiencing homelessness or were vulnerably housed at the time of enrolment. Participants were referred to the study by clinical staff through Ottawa Inner City Health and the Royal Ottawa Hospital’s Psychiatric Outreach Team. Ottawa Inner City Health provides comprehensive health services, including mental health care to Ottawa’s homeless community. The Psychiatric Outreach Team is a community-based short-term service providing support and referrals to individuals who are homeless or vulnerably housed experiencing severe and persistent mental illness. Eligibility criteria are described in Figure 1.

**Figure 1.**
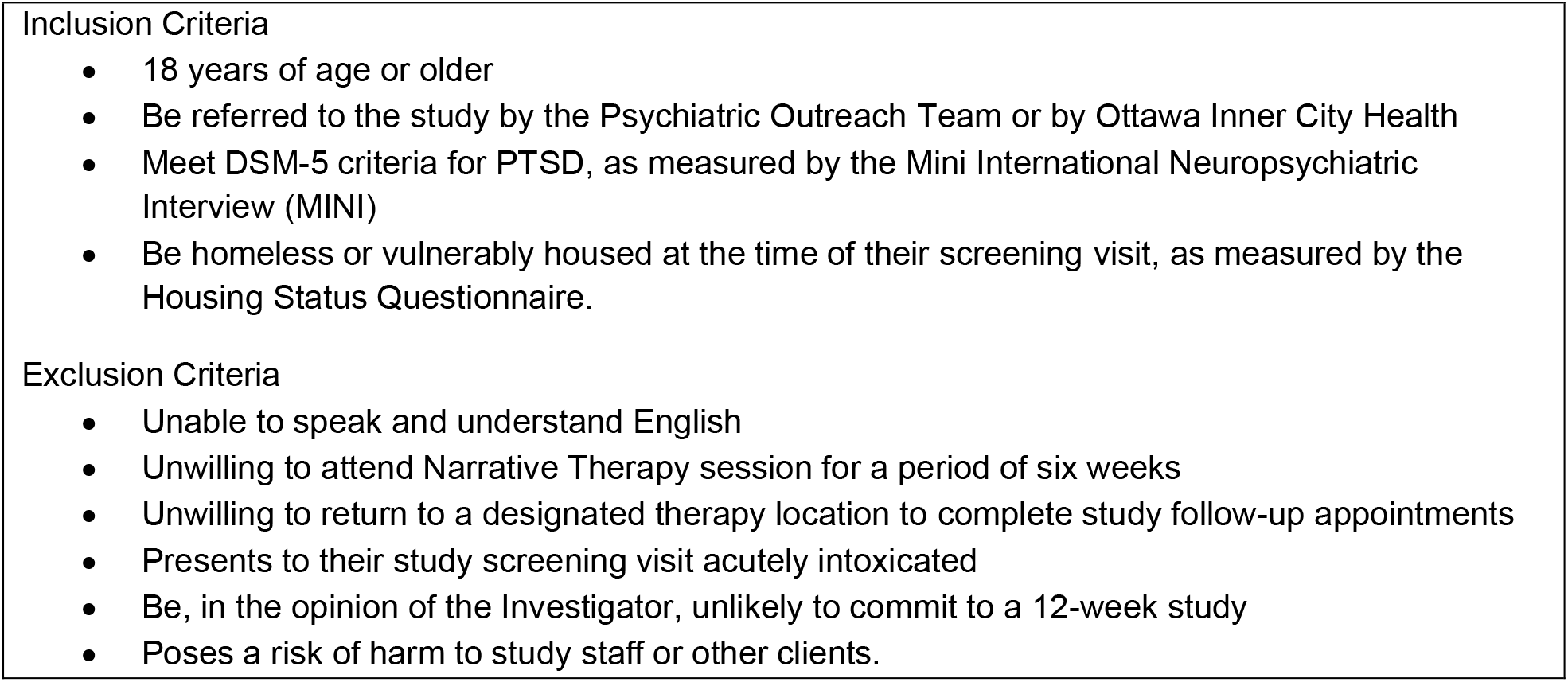
Participant Eligibility Criteria.

### Interventions

Participants were randomized to one of two groups: Narrative Exposure Therapy only (NET) or Narrative Exposure Therapy supplemented with a genealogist assessment (NET+G). Two therapists, a psychiatrist [SH] and a social worker [MSW – KB], who had received specialist training in NET, provided the therapy in the study. NET was delivered as described by Schauer et al(29). The components of NET as delivered in this study are outlined in Table 1. Participants attended weekly visits with the therapist at the location of their choosing. Flexibility in timing was offered to participants who found it challenging to attend weekly visits. Participants randomized to the NET+G arm were connected to a genealogist [MG] after their baseline visit to complete a family history interview and voluntary genetic testing. Participants were provided with a full family history report detailing any information discovered through the interview and matching with public genealogy databases. For this study, Family Tree DNA was used for genetic matching.

**Table 1.** Components of Narrative Exposure Therapy.

|  |  |
| --- | --- |
| Part 1 –<br>Session 1 | Completion of structured diagnostic measures (CAPS-5, PCL-5 with LEC and Criterion A)<br>Psychoeducation and outline of the therapeutic plan |
| Part 2 –<br>Session 2 | Construction of the Lifeline – a visual biographical overview of significant life events |
| Part 3 –<br>Sessions 3-6 | *Sessions 3 -5*<br>Narration of the lifeline from birth through each event<br>• Traumatic events are confronted and reprocessed until arousal response decreases<br>• At follow-up sessions, the draft narrative is read through collaboratively, focusing on important events.<br>• The narrative is updated and corrected, providing more clarity with each read-through<br>• This is repeated until a final version of the narrative is completed (by Session 6)<br>*Session 6*<br>• The final narrative is read through entirely and signed by the participant and therapist.<br>• The participant is provided a copy of their narrative |

*Adapted from Schauer et al 2011.*

Participants were met in the community at a mutually agreed upon location. Visit locations included shelters, outreach offices, community day programs and our research office. Locations were chosen to minimize participant burden and increase feelings of comfort and safety. Any required travel costs were covered by the study.

After referral to the study, participants met with a trained research assistant to complete the informed consent process and screening procedures. Screening procedures to confirm eligibility involved a structured interview completed by the research assistant to confirm active symptoms of PTSD and housing status. Once eligibility was confirmed, participants completed assessments collecting demographics and evaluating general mental health, alcohol and substance use, quality of life, health care utilization, and cognitive state. We collected sociodemographic information on gender (male, female, transgender, non-binary), self-identified ethnicity, highest level of education completed, marital status and medical history.

### Outcomes

The primary outcomes of this study were the feasibility and acceptability of a completing an RCT of NET in the homeless population. The primary measure of feasibility was recruiting the planned sample size over six months. In addition, we wanted to determine the acceptability of NET in this population which we defined as at least 50% of those approached about the study consenting to take part. Lastly, we wanted to see if it was feasible to collect outcome data in this population which we defined as restricting study dropouts or lost to follow ups to 25% of participants.

We also wanted to see if NET treatment in this population resulted in better health-related outcomes including: a decrease in the severity of PTSD symptoms, change in housing status, improved overall health, better health-related quality of life and lower rates of alcohol/drug misuse. Other secondary outcomes included creating a training manual for Narrative Exposure Therapy in this population that also included the incorporation of a genealogist.

Measures and timing of administration are outlined in Table 2. Housing status was collected prior to entry into the study and at each follow-up visit. Participants self-identified their housing among the following options: living in a shelter, living with a friend, living with family, supportive/transitional housing, paying for a space, no housing options, or other. Participants could select more than one option if applicable. The Addiction Severity Index interview(40) was completed at baseline, weeks 4, 8 and 12 to evaluate both lifetime substance use and use in the past 30 days. We calculated the total number of days of use in the past 30 days for each time point to evaluate any changes in drug or alcohol use between visits. The SF-20 was used from February 2019 until September 2019, when it was replaced with the EQ-5D-5L. The decision was made to change measures as participants reported substantial difficulty in completing the SF-20 and interpreting the questions. The EQ-5D-5L is much shorter, with only 5 questions plus the visual analogue scale, and was much better tolerated as a measure. Participants who started with the SF-20 continued to use it through their follow up time points.

**Table 2.** Time and Events Schedule.

| Visit/Measure | Measure Description | Screening/<br>Baseline | Week 1 | Week 4 | Week 8<br>Follow-up | Week 12<br>Follow-up |
| --- | --- | --- | --- | --- | --- | --- |
| Housing Status | A brief self-reported measuring describing participant's housing status | X | - | X | X | X |
| MINI – PTSD Module | A structured interview assessing symptoms of PTSD in the past 30 days. | X | - | - | - | - |
| CAPS-5 | A structured interview, administered during therapy, to assess a traumatic event(s) and associated symptoms in the past 30 days | - | X | - | - | - |
| PCL-5 with Life Events Checklist | A 20 item self-report evaluating the severity of PTSD symptoms over the previous 30 days; LEC assesses exposure to 16 events known to potentially result in PTSD | - | X | - | X<br>(PCL-5 only) | X<br>(PCL-5 only) |
| Demographics | Self-reported sociodemographic variables including gender, sexual orientation, ethnicity, education, and marital status | X | - | - | - | - |
| Addiction Severity Index | A semi-structured interview used to assess severity of substance abuse problems, both lifetime and past 30 days | X | - | X | X | X |
| AUDIT | A brief 10-item self report questionnaire assessing alcohol misuse | X | - | X | X | X |
| ADHD Self-Report Scale | An 18-item self-report questionnaire that assesses the inattentive (6 items) and hyperactive/impulsive (12 items) dimensions of ADHD | X | - | - | - | - |
| Montreal Cognitive Assessment | A brief interview assessing multiple cognitive domains including visuo-constructional skills; naming; memory; attention; sentence repetition; verbal fluency; abstraction; delayed recall; and, orientation. | X | - | - | - | - |
| Short Form Health Survey (SF-20) or EQ-5D-5L | The SF-20 was used from February 2019 – September 2019. This is a 20-item questionnaire that assesses various health outcomes, and the extent to which health related problems interfere with daily life. | X | - | X | X | X |
|  | The EQ-5D-5L was used from September 2019 – end of study. The EQ-5D-5L is 5-item questionnaire that assesses health-related quality of life. |  |  |  |  |  |
| Health Care Costs | A self-report questionnaire asking about missed time from work/volunteer/school and utilization of healthcare services. | X | - | X | X | X |

### Sample Size

We planned to enroll 12 participants in each arm for a total sample of 24 participants(41). The study was not powered to test efficacy outcomes.

### Randomization and blinding

Randomization was completed by the Ottawa Methods Centre at the Ottawa Hospital Research Institute (OHRI) with allocations kept in sequential sealed opaque envelopes at the OHRI study office. Participants were randomized in a 1:1 allocation with no restrictions. After providing consent and confirming eligibility, participants were randomized by a trained research assistant according to the allocation in the sealed envelope.

### Statistical methods

All statistical analyses were conducted using IBM SPSS 26. Non-parametric data were described using frequencies and percentages. Continuous data were described using measures of central tendency (mean and standard deviation) and bivariate relationships were explored using independent samples t-tests (t) and analyses of variance (ANOVA). Paired samples t-tests were conducted to explore any within subject relationships for PCL-5 score and substance use patterns. Bivariate relationships were detected using two-sided Fisher’s Exact Test (2×2 contingency tables), except for one-sided tests where noted in the tables.

### Qualitative Evaluation

Qualitative interviews were planned with participants who completed the study and service providers including shelter, outreach, and day program staff. The service provider interview evaluated their participation in the study, their perceived need for community-based NET to be implemented in this population and what supports might be necessary to implement either a large-scale trial or a permanent service offering NET to individuals who are homeless or vulnerably housed.

#### Ethics and Informed Consent

Research ethics approval was provided by the Royal Ottawa’s Institute of Mental Health Research Ethics Board (IMHR-REB ID: 2017042) and the Ottawa Health Sciences Network REB (OHSN-REB ID: 20180895-01H). Written informed consent was obtained from the participants who were completely informed of the study purposes and procedures.

## Results

### Participant flow

Participant flow is outlined in Figure 2. Recruitment for this study took place between February 2019 and February 2020 and a total of 22 people were referred to the study, (one of these was immediately before the first pandemic lockdown in late February 2020). Recruitment was stopped on two occasions, for a total of three months, as therapists had reached their maximum case load. Five participants were referred to the study after the start of the COVID-19 pandemic in March 2020, but we were unable to recruit them as there were no safe places to meet face to face as most drop-in centres or other spaces were closed because of COVID-19. We have not included them in the tables. Virtual appointments were not an option for this population due to a lack of technology, access to the internet, or safe and private places to conduct therapy.

**Figure 2.**
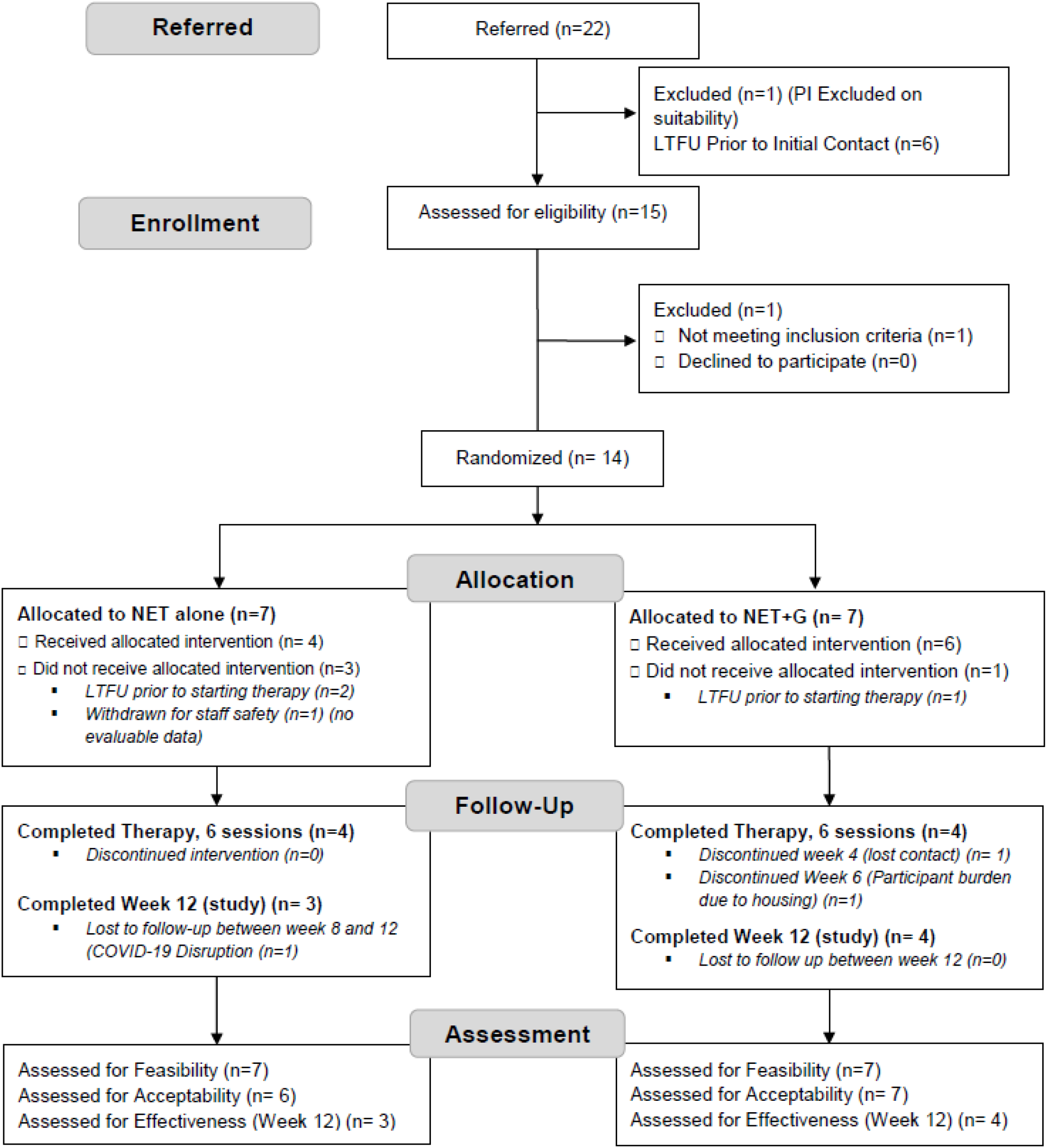
Consolidated Standards of Reporting Trials flow and attrition diagram.

The clinical teams referred 22 people to take part in the study over the nine months that the study was accepting potential participants. One potential participant was deemed ineligible by the Principal Investigator due to a significant brain injury and associated psychosis. Six (6/22, 27%) potential participants could not be contacted to arrange a baseline visit and consent, leaving 15/22 (68%) who were assessed for eligibility for the study. One of these potential participants did not meet the inclusion criteria. This participant consented but was unable to complete the screening procedures. This led the team to evaluate the administration of screening measures to reduce burden on participants.

This left 14 participants who were randomized. Seven were randomised to NET and seven to NET+G. One participant who consented to the study and was randomized to the NET alone arm did not complete their baseline assessment. While consent, screening and baseline typically occurred at one visit, this participant had a particularly long screening session, and the baseline assessments were scheduled to occur during a second encounter. The participant did not present for this visit and further information from the referrer showed a possible safety risk to the therapists, so this potential participant was withdrawn. This participant had no evaluable data and was not included in the evaluation of acceptability or effectiveness.

Three people were lost to follow-up between consenting to take part in the study and the first therapy session, one in the NET+G group and two in the NET group.

Every effort was made to provide participants with both their personal narrative and genealogy report before their final NET session. In particular, delays with genetic matching meant the first participant in the NET+G arm did not receive their report until after their final session. Adjustments were made in the timing of the genealogy interview so that all subsequent participants received their reports prior to completing therapy, and typically around week 4.

### Feasibility

We were unable to recruit our desired sample size within the goal of 6 months. The enrollment of 15 participants took place over approximately 9 months. This was primarily due to a shortage of trained therapists to take on participants. For the first 5 months, only one therapist was involved in the study. As the study progressed, we were able to recruit one additional trained therapist.

### Study Acceptability

Figure 2 outlines the retention rates at various stages of the study. While we did not meet the a priori threshold of 75% retention at week 12, 61.5% (8/13) of participants completed at least one post-therapy follow-up assessment visit (week 8). Of the ten who started therapy, seven completed all study visits up to week 12. Of three participants who did not complete the week 12 assessment, the first completed the week 8 study visit and was interested in completing week 12, however, the final visit was rescheduled due to personal circumstances and was eventually cancelled due to the COVID-19 pandemic; the second dropped out at week 6 as they found the sessions too triggering and difficult to complete while sleeping rough; and, the third individual was lost to follow up at week 4 but re-engaged with the clinical team at a later date.

### Therapy Acceptability

Of the 10 participants who started therapy, eight (80%) completed all 6 sessions. All participants who were randomized to the NET+G arm (n=6) accepted the referral to complete their family history.

### Demographics

The demographics of participants are outlined in Table 3. Comparable to community demographics, our sample contained more males than females (61.5%, 8/13), with a lifetime history of drug use (84.6%, 11/13) and alcohol use (84.6%, 11/13). The sample self-identified primarily as white (65%, 8/13), but also First Nations (7.7%, 1/13), Métis (2/13, 15.4%), Asian (7.7%, 1/13), and Other (Hispanic) (7.7%, 1/13). Within this identification, two people also identified themselves as mixed-ethnicity (Métis/White and First Nations/Black). Educational background was diverse with 38.5% (5/13) having less than high school, 30.8% (4/13) having a high school diploma or equivalent, 15.4% (2/13) with a college or university education, and 15.4% (2/13) with a graduate or professional degree. Marital status was also varied, with 46.2% (6/13) identifying as single, 7.7% (1/13) as married, 23.1% (3/13) as separated, 7.7% (1/13) as divorced and 15.4% (2/13) as widowed. No individuals identified as transgender or non-binary. No participants indicated common-law status.

**Table 3.**
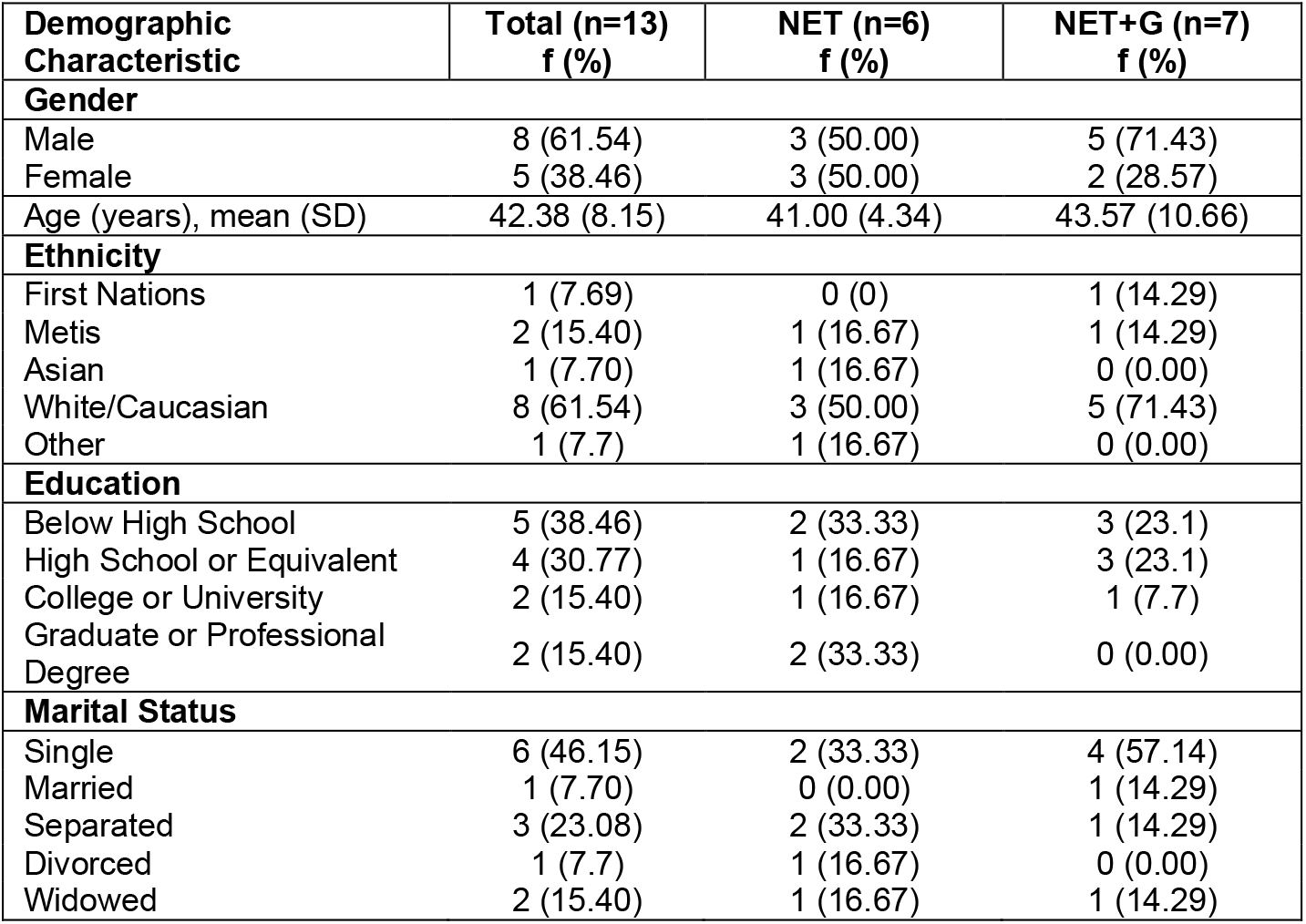
Demographic Characteristics.

### PTSD Scores

All participants had experienced multiple traumatic events in their lifetime, with childhood trauma being common, as reported on the Life Events Checklist. With respect to symptom severity, baseline PCL-5 scores did not differ (*F*(1,13)=1.169,p=.311) between those allocated to NET (M=66.00, SD=6.68) or NET+G (M=60.67, SD=8.17).

PTSD scores decreased in both groups over the course of the study. Assessing the change in symptom severity within subjects for the 7 who completed post-therapy follow-up to week 12, showed a statistically significant improvement in PTSD scores (Table 4). Prior to initiating therapy, the average PCL-5 score was 64.14 (SD 8.80). At the 12 week follow-up, participants reported an average decrease of 17.29 points (SD 6.18), for a total PCL-5 score of 46.86 (SD 16.63) (*t*=2.798, *df*=6, p=.031, *d*=1.057). This reduction in severity within subjects is also considered a clinically meaningful change, defined as a reduction in total score by 10-20 points(42).

**Table 4.** Within subject outcomes.

| Time Point | PCL-5 Scores<br>(n=7) |  |  | Drug Use (Previous 30 Days)<br>(n=7) |  |  | Alcohol Use (Previous 30 Days)<br>(n=7) |  |  |
| --- | --- | --- | --- | --- | --- | --- | --- | --- | --- |
|  | M | SD | Paired Samples t-test | M | SD | Paired Samples t-test | M | SD | Paired Samples t-test |
| Week 0 | 64.14 | 8.80 | $t= 2.798$<br>$df\ 6,$<br>$p= .031$<br>$d=1.057$ | 11.57 | 14.74 | $t= 1.00$<br>$df\ 6,$<br>$p= .356$ | 1.00 | 1.73 | $t= 1.08$<br>$df\ 6,$<br>$p= .321$ |
| Week 12 | 46.86 | 16.63 |  | 10.71 | 14.27 |  | 0.43 | 0.787 |  |

### Substance Use

A lifetime history of substance use was common in this sample, with 84.6% (11/13) reporting a history of drug or alcohol misuse. At enrolment, 6 individuals reported current drug use (46.2%, 6/13) and 2 reported alcohol use (15.4%, 2/13) in the 30 days prior, while 5 participants reported no drug or alcohol use at all during this period (38.5%, 5/13). Alcohol or drug use did not change over the duration of the study (Table 4). Prior to initiating therapy, drug use was an average of 11.57 days (SD 14.47 days), while alcohol use averaged 1 day (SD 1.73 days). At week 12, there was no significant decrease in drug use (*t=1*.*00, df= 6*, p=.356) or alcohol use (*t*=1.08, *df*=6, p=.321).

Substance use appeared to have no relationship to whether or not a participant completed the study, with 23.1% of participants using drugs at enrolment completing the study (p=.617, Fisher’s Exact Test, 1-tailed) compared to 30.7% of non-users completing the study. Participants who did not complete the study were split, with 23.1% using drugs at enrolment and 23.1% not using drugs (Table 5).

**Table 5.** Study completion status.

|  |  | **Baseline**<br>**N=13** |  |
| --- | --- | --- | --- |
|  |  | Study Complete<br>f (%) | Study Not Complete<br>f (%) |
| Drug Use<br>(Past 30 Days) | Yes | 3/13 (23.1) | 3/13 (23.1) |
|  | No | 4/13 (30.7) | 3/13 (23.1) |
|  | *Fisher's Exact Test* | p= .617 (1-tailed) |  |
| Alcohol Use<br>(Past 30) | Yes | 2/13 (15.4) | 0/13 (0) |
|  | No | 5/13 (38.5) | 6/13 (46.1) |
|  | *Fisher's Exact Test* | p= .269 (1-tailed) |  |
| Housing Status | Shelter/Rough | 2/13 (15.4) | 5/13 (38.5) |
|  | Other | 5/13 (38.5) | 1/13 (7.6) |
|  | *Fisher's Exact Test* | p= .078 (1-tailed) |  |

Similarly, no statistically significant difference was found between those who completed the study and were using alcohol (2/13, 15.4%) or not using alcohol (5/13, 38.5%) compared to the six participants not using alcohol (6/13, 46.2%) who did not complete the study (p=.269, Fisher’s Exact Test, 1-tailed) (Table 5).

### Housing Status

Housing status over the duration of the study is described in Table 6. At baseline, more than half the participants were living in a shelter (53.8%, 7/13), followed by supportive/transitional housing (15.4%, 2/13), while 1 individual was living with a friend (7.7%), and another was paying for a living space (7.7%). Several individuals added specifiers to their situation including that they were paying for a living space because they received a subsidy or assistance from a family member. During the study, one individual (7.7%) left the shelter and was not able to find other housing options. Of those who did not complete the study, 38.5% (5/13) resided in a shelter baseline (p=.078, Fisher’s Exact Test, 1-tailed).

**Table 6.** Housing status.

| Housing Status (N=13) | Baseline | Week 4 | Week 8 | Week 12 |
| --- | --- | --- | --- | --- |
|  | f (%) |  |  |  |
| Shelter | 7 (53.8) | 3 (23.1) | 4 (30.8) | 3 (23.1) |
| With Family | 1 (7.7) | 1 (7.7) | 1 (7.7) | 1 (7.7) |
| With Friend | 0 (0) | 0 (0) | 0 (0) | 1 (7.7) |
| Supportive/Transitional Housing | 4 (30.8) | 2 (15.4) | 1 (7.7) | 1 (7.7) |
| Paying for a Space | 1 (7.7) | 1 (7.7) | 2 (15.4) | 1 (7.7) |
| No Housing Options | 0 (0) | 1 (7.7) | 0 (0) | 0 (0) |
| Missing | 0 (0) | 1 (7.7) | 0 (0) | 0 (0) |
| Withdrawn | 0 (0) | 4 (30.8) | 5 (38.5) | 6 (46.2) |

Of those who completed the study, 3 individuals remained at the shelter (3/7), while others were staying with friends (1/7), staying with family (1/7), living in supportive housing (1/7), or paying for a space (1/7). Several individuals experienced changes to their housing situation: two moved from supportive living to the shelter, one from the shelter to living with a friend, one from supportive housing to paying for a space, one from living with family to paying for a living space. Of the 6 individuals who did not complete the study, 4 were at the shelter, 1 was “sleeping rough”, and 1 was paying for a living space.

#### Qualitative Evaluation

We were unable to complete qualitative interviews with the participants because of COVID-19 restrictions. There were no safe places to conduct interviews and using technology was not feasible as discussed above. Four service providers completed a virtual semi-structured qualitative interview with an unfamiliar research staff member. The service providers had varying backgrounds including two staff with Psychiatric Outreach who referred clients to the study, one manager at a shelter, and one manager at a community day program, both of whom facilitated study activities.

### Study Participation

Study providers identified no difficulties in communicating with the study team. Generally, expectations were met with respect to each individual’s anticipated role in the study. One service provider identified that additional guidance on who might be a good candidate and what role they need to play after enrolment in the study would be beneficial. All four service providers felt that both study participation and the delivery of NET integrated well with the services that they currently offer. Clinical providers felt that it was beneficial to have resources that they could offer these clients, in particular noting the benefit to clients not having to attend the hospital for care.

### Need for a Trauma Therapy

The service providers unanimously expressed the need for this service within their community. One provider when asked about the need for NET as a service noted: “It’s just more than needed, it’s irreplaceable, without it there’s nothing. Nothing, like, is really happening right now so it would be great to implement this in a community setting on a broader scale.” Similarly, a second provider noted that NET was “absolutely necessary, nothing to really add, there needs to be this type of service for this population, the challenge isn’t figuring out if there’s a need but more how can we provide what’s needed so definitely a need.”

### Implementation of NET as a Service

The service providers were supportive of both the conduct of a large-scale trial all expressed interest in participating in such an initiative. With respect to how to offer NET on a larger scale, the lack of resources, both time and trained staff, was a common concern, however they were all amenable to a centralized resource as an option to delivery therapy services in the community as well. Despite the potential staffing constraint, the service providers indicated their willingness to navigate any barriers that may exist to continuing to offer this therapy in their respective settings. All indicated interest in having a role in further research studies and were keen to facilitate this research.

## Discussion

### Generalizability

The sample enrolled in the study is largely representative of the homeless population, both in Ottawa and nationally. It is reasonable to assume that our results may be generalizable and valid across other homeless populations. However, it would be worthwhile conducting a large RCT in multiple communities to increase diversity and the inclusion of people experiencing homelessness in smaller centres.

### Interpretation

Individuals experiencing homelessness have long been excluded from clinical trials with the assumption that they would be challenging to engage or retain. We found that, while not quite meeting the pre-defined thresholds for feasibility and acceptability, conducting a RCT of a mental health intervention in the community was both feasible and acceptable. To improve feasibility and acceptability during a large scale RCT, several lessons learned from this pilot can be used to improve both engagement and retention.

A lack of therapists trained in NET had the largest impact on meeting our feasibility target of recruiting 24 individuals within 6 months. The COVID-19 pandemic also impacted recruitment with no individuals enrolled after February 2020. NET training is limited with only 2 offerings occurring in the Ottawa area within the 3-year study period. The Principal Investigator [SH] recently organized two virtual training session with high levels of community interest. Building therapist capacity will be critical to the success of implementing a large RCT and in offering NET as a standard of care long term in the community. The provision of NET by health professional students supervised by experienced therapists is also a model that needs to be explored especially given most universities’ commitment to social accountability in their communities.

While our initial target of 75% retention was not met, our retention rate is consistent with literature around engagement in psychotherapy, while our proportion of participants for completing all sessions of therapy (80%) exceeded average retention rates(43,44). Given the considerations of working with individuals who are homeless and the potential challenges of engaging with trauma therapy, we feel that these rates are high enough to deem the therapy and participation in a study to be acceptable.

What is striking is that the drop-out rate before starting therapy is much higher than after starting treatment, with12/22 (55%) of potential participants dropping out prior to therapy compared to only two out of ten (20%) after starting therapy. It is likely that this is due to a combination of three factors. First is that referrers to the study need to be given clear guidance as to who is potentially eligible and who is not. Also providing information about what happens after therapy is completed would be helpful. Second, there are real practical difficulties in contacting people who are vulnerably housed in a way and at a time which is convenient to them. Third, there are the issues of credibility and trust. People who have experienced trauma, especially in childhood, have learnt not to trust carers and many may have had bad experiences in their contacts with the health system. The contractual process of signing an informed consent form is probably not sufficient to gain trust without other actions. These could include paying attention to privacy of data, linking the study to creditable organizations and being clear about who has what role in the research team and how long this will last. Requiring potential participants to see several unfamiliar people before starting therapy may also impact on trust. Training research assistants in helping people with complex PTSD and how it impacts on relationships is important. This would be guided by trauma informed research practices, such work done by Voith and colleagues(45), and would seem to be an important component of a bigger trial.

Substance use at the time of enrolment did not appear to impact whether or not a participant was likely to complete therapy and follow-up sessions, suggesting that it is feasible to include those using substances in research and therapy. However, staying in a shelter at the time of enrolment did approach significance with respect to not completing the study. Given this, consideration should be given when working with individuals who are staying in a shelter on how to best support therapy retention. Factors to increase retention should be examined during the optimization phase of MOST, with a particular focus on those who are in the shelter or sleeping rough.

Within subject differences showed a significant and clinically meaningful reduction in PTSD symptoms in participants from baseline to week 12, suggesting that Narrative Exposure Therapy may be effective in this population. Further research through a large scale RCT is needed to determine the effect of genealogy and optimize the delivery of NET in this population.

### Strengths and limitations

The major strength of this study is that we have shown that it is both feasible and acceptable to conduct an RCT for the treatment of trauma with individuals experiencing homelessness in a community setting – the first study of its kind. Additionally, the study also shows preliminary evidence for NET as an effective therapy for complex trauma in the homeless. However, the study is not without limitations. The major limitation of this study was a lack of individuals trained in NET. As a pilot feasibility study, the trial is also underpowered to detect any significant differences between treatment groups.

With respect to the genealogy report, it took approximately 4-6 weeks to receive a completed report, which made it challenging to incorporate the findings of the report into the narrative process. It would be possible in a large-scale trial to ensure that the individual is seen by the genealogist in advance of initiating therapy to increase the likelihood of receiving the report during active narration.

Lastly, due to the sudden lockdown measures implemented during the COVID-19 crisis the study team was unable to complete the planned qualitative interviews about participant experiences in the study, the therapy or genealogy support. Anecdotally, there were some complaints about the complexity of two questionnaires (SF-20 and Health Care Costs Questionnaire), but participants did not refuse to answer any questionnaires or comment on the length of visits. With respect to the genealogist, participants all received their reports, with one participant lost to follow-up requesting their report several months later. Only one participant, at the time of consent, indicated that they would have no interest in speaking with a genealogist if they were randomized to that arm.

### Clinical Considerations

There is currently a significant gap in trauma treatment for the homeless community, despite the high prevalence and degree of complexity within this population. This study has shown that not only is it feasible to deliver community-based therapy without a fixed location, but that Narrative Exposure Therapy is an effective and acceptable therapeutic option for individuals experiencing complex trauma and homelessness. Additionally, service providers have emphatically endorsed the need for this program after their participation in the study.

### Conclusions and Future Research

This study was the first of three components of the preparation stage of a multiphase optimization strategy (MOST). The second is a scoping review (46) which we have published separately on the treatment of PTSD in the homeless including the use of trauma informed care to deliver such therapy. The third is a consultation with key stakeholders, including individuals with lived experience, about the implementation of a trauma informed care model for individuals who are homeless or vulnerably housed. A focus of this consultation has been addressing the issue of trust when referring potential participants to a treatment study. We will use these components to develop a model of delivering NET in this population which we will optimize using a factorial study in preparation for a larger multi-site RCT to evaluate the efficacy of NET as well as genealogy support in this population. The multi-site RCT will also help to evaluate issues of generalizability across communities that have different supports for their homeless population.

## Data Availability

The datasets used and/or analyzed during the current study are available from the corresponding author on reasonable request.

## List of Abbreviations

NET: Narrative Exposure Therapy
RCT: Randomized Controlled Trial
PTSD: Post-Traumatic Stress Disorder
cPTSD: Complex Post-Traumatic Stress Disorder
MOST: Multi-phase Optimization Strategy
NET+G: Narrative Exposure Therapy Plus Genealogy
OHRI: Ottawa Hospital Research Institute

## Declarations

### Ethics

Research ethics approval, including consent procedures, was provided by the Royal Ottawa’s Institute of Mental Health Research Ethics Board (IMHR-REB ID: 2017042) and the Ottawa Health Sciences Network REB (OHSN-REB ID: 20180895-01H).

### Consent for Publication

Not applicable.

### Author Contributions

NEE was involved in protocol development, study conduct, data analysis and was a major contributor in writing the manuscript. AB was involved in study conduct. NSD was involved in study design, protocol development, and study conduct. SEM was involved in study design and SH was involved in obtaining funding, study design, protocol development, conducting therapy sessions, data analysis and interpretation, and was a major contributor in writing the manuscript. All authors read and approved the final manuscript.

## Acknowledgements

The study team would also like to recognize the following individuals for their contributions to the project: Mags Gaulden (genealogist), Kim Bulger (therapist), research staff Brooklyn Ward, and the service users who both participated and advised on the design and conduct of this study. The study team would also like to thank the Royal Ottawa’s Psychiatric Outreach Program, Ottawa Inner City Health, Centre 454, and the Ottawa Salvation Army (Booth Street) for their administrative support of the trial.

## References

1. Canadian Observatory on Homelessness. How many people are homeless in Canada? [Internet]. 2013 [cited 2021 Sep 21]. Available from: https://www.homelesshub.ca/about-homelessness/homelessness-101/how-many-people-are-homeless-canada

2. Employment and Social Development Canada. Everyone Counts 2018: Highlights: Preliminary results from the second nationally coordinated Point-in-Time count of homelessness in Canadian communities [Internet]. 2018. Available from: https://www.canada.ca/content/dam/esdc-edsc/documents/programs/homelessness/reports/1981-Reaching_Home-PIT-EN_(3).pdf

3. Kingston Frontenac Lennox Addington United Way. Results of the Urban Kingston Point-in-Time Count [Internet]. 2021. Available from: https://www.unitedwaykfla.ca/wp-content/uploads/2021/09/2021-Kingston-PiT-Count-Results.pdf

4. Niagra Region Homelessness Services. Homelessness Point-in-Time Count Report - Niagra Region. 2021.

5. US Department of Housing and Urban Development. The 2020 Annual Homeless Assessment Report to Congress. Development. 2020;

6. Hwang SW. Homelessness and health. CMAJ. 2001.

7. Omerov P, Craftman ÅG, Mattsson E, Klarare A. Homeless persons’ experiences of health-and social care: A systematic integrative review. Health Soc Care Community [Internet]. 2020 Jan 16 [cited 2021 Feb 8];28(1):1–11. Available from: https://onlinelibrary.wiley.com/doi/abs/10.1111/hsc.12857

8. Hwang SW, Aubry T, Palepu A, Farrell S, Nisenbaum R, Hubley AM, et al. The health and housing in transition study: A longitudinal study of the health of homeless and vulnerably housed adults in three Canadian cities. Int J Public Health. 2011;

9. Purkey E, MacKenzie M. Experience of healthcare among the homeless and vulnerably housed a qualitative study: Opportunities for equity-oriented health care. Int J Equity Health. 2019;

10. Gilmer C, Buccieri K. Homeless Patients Associate Clinician Bias With Suboptimal Care for Mental Illness, Addictions, and Chronic Pain. J Prim Care Community Heal. 2020;

11. Wen CK, Hudak PL, Hwang SW. Homeless people’s perceptions of welcomeness and unwelcomeness in healthcare encounters. J Gen Intern Med. 2007;

12. McInerney SJ, Fallahi A, Edgar NE, Ceniti AK, Rizvi SJ, Beder M, et al. Suicide-related presentations of homeless individuals to an inner-city emergency department. General Hospital Psychiatry. 2020.

13. Anderson I, Ytrehus S. Re-conceptualising approaches to meeting the health needs of homeless people. J Soc Policy. 2012;

14. Buhrich N, Hodder T, Teesson M. Lifetime Prevalence of Trauma among Homeless People in Sydney. Aust New Zeal J Psychiatry [Internet]. 2000 Dec 21 [cited 2020 Nov 8];34(6):963–6. Available from: http://journals.sagepub.com/doi/10.1080/000486700270

15. Keane CA, Magee CA, Kelly PJ. Is there Complex Trauma Experience typology for Australian’s experiencing extreme social disadvantage and low housing stability? Child Abus Negl. 2016 Nov 1;61:43–54.

16. Taylor KM, Sharpe L. Trauma and post-traumatic stress disorder among homeless adults in Sydney. Aust N Z J Psychiatry [Internet]. 2008 Mar 1 [cited 2020 Nov 8];42(3):206–13. Available from: http://journals.sagepub.com/doi/10.1080/00048670701827218

17. Baggett TP, O’Connell JJ, Singer DE, Rigotti NA. The unmet health care needs of homeless adults: A national study. Am J Public Health [Internet]. 2010 Jul 1 [cited 2020 Nov 8];100(7):1326–33. Available from: http://ajph.aphapublications.org/doi/10.2105/AJPH.2009.180109

18. Carlson EB, Garvert DW, Macia KS, Ruzek JI, Burling TA. Traumatic Stressor Exposure and Post-Traumatic Symptoms in Homeless Veterans. Mil Med [Internet]. 2013 Sep 1 [cited 2020 Nov 8];178(9):970–3. Available from: https://academic.oup.com/milmed/article/178/9/970-973/4259610

19. Whitbeck LB, Armenta BE, Gentzler KC. Homelessness-Related Traumatic Events and PTSD Among Women Experiencing Episodes of Homelessness in Three U.S. Cities. J Trauma Stress [Internet]. 2015 Aug 1 [cited 2020 Nov 8];28(4):355–60. Available from: http://doi.wiley.com/10.1002/jts.22024

20. Torchalla I, Strehlau V, Li K, Linden IA, Noel F, Krausz M. Posttraumatic stress disorder and substance use disorder comorbidity in homeless adults: Prevalence, correlates, and sex differences. Psychol Addict Behav [Internet]. 2014 [cited 2020 Nov 8];28(2):443–52. Available from: /record/2013-27999-001

21. Van Ameringen M, Mancini C, Patterson B, Boyle MH. Post-traumatic stress disorder in Canada. CNS Neurosci Ther. 2008 Sep;14(3):171–81.

22. Goodman L, Saxe L, Harvey M. Homelessness as psychological trauma: Broadening perspectives. Am Psychol. 1991;

23. Hopper EK, Bassuk EL, Olivet J. Shelter from the storm: Trauma-informed care in homelessness services settings. Open Health Serv Policy J [Internet]. 2009;2:131–51. Available from: http://www.traumacenter.org/products/pdf_files/shelter_from_storm.pdf

24. World Health Organization. International Classification of Diseases for Mortalility and Morbidity Statistics Eleventh Edition. World Health Organization. 2021.

25. Armstrong R, Phillips L, Alkemade N, Louise O’Donnell M. Using latent class analysis to support the ICD-11 complex posttraumatic stress disorder diagnosis in a sample of homeless adults. J Trauma Stress. 2020 Oct 1;33(5):677–87.

26. Møller L, Augsburger M, Elklit A, Søgaard U, Simonsen E. Traumatic experiences, ICD-11 PTSD, ICD-11 complex PTSD, and the overlap with ICD-10 diagnoses. Acta Psychiatr Scand. 2020;

27. Magwood O, Leki VY, Kpade V, Saad A, Alkhateeb Q, Gebremeskel A, et al. Common trust and personal safety issues: A systematic review on the acceptability of health and social interventions for persons with lived experience of homelessness. PLoS One. 2019 Dec 1;14(12):e0226306.

28. Hwang SW, Ueng JJM, Chiu S, Kiss A, Tolomiczenko G, Cowan L, et al. Universal health insurance and health care access for homeless persons. Am J Public Health. 2010;

29. Schauer M, Elbert T, Neuner F. Narrative exposure therapy : A short-term treatment for traumatic stress disorders. Hogrefe; 2011. 110 p.

30. American Psychological Association. Clinical Practice Guideline for the Treatment of Posttraumatic Stress Disorder (PTSD). Washington, DC APA, Guidel Dev Panel Treat Posttraumatic Stress Disord Adults. 2017;

31. NICE. Post-traumatic stress disorder: NICE guideline. NICE Guidance. 2018.

32. Crombach A, Elbert T. Controlling Offensive Behavior Using Narrative Exposure Therapy: A Randomized Controlled Trial of Former Street Children. Clin Psychol Sci. 2015;

33. Mørkved N, Hartmann K, Aarsheim LM, Holen D, Milde AM, Bomyea J, et al. A comparison of narrative exposure therapy and prolonged exposure therapy for PTSD. Clinical Psychology Review. 2014.

34. Bowen P, Rose R, Pilkington2 A. Mixed Methods-Theory and practise.Sequential, explanatory approach. International Journal of Quantitative and Qualitative Research Methods. 2017.

35. Goodman RD. The transgenerational trauma and resilience genogram. Couns Psychol Q [Internet]. 2013 Dec [cited 2021 Mar 3];26(3–4):386–405. Available from: https://www.tandfonline.com/doi/abs/10.1080/09515070.2013.820172

36. Mitchell MD, Shillingford MA. A Journey to the Past. Fam J [Internet]. 2017 Jan 21 [cited 2021 Mar 3];25(1):63–9. Available from: http://journals.sagepub.com/doi/10.1177/1066480716679656

37. Mahuika N, Kukutai T. Introduction: Indigenous Perspectives on Genealogical Research. Genealogy. 2021;

38. Hatcher S, Coupe N, Wikiriwhi K, Durie SM, Pillai A. Te Ira Tangata: a Zelen randomised controlled trial of a culturally informed treatment compared to treatment as usual in Māori who present to hospital after self-harm. Soc Psychiatry Psychiatr Epidemiol. 2016;

39. Eldridge SM, Chan CL, Campbell MJ, Bond CM, Hopewell S, Thabane L, et al. CONSORT 2010 statement: Extension to randomised pilot and feasibility trials. Pilot Feasibility Stud. 2016;

40. Zanis DA, McLellan AT, Cnaan RA, Randall M. Reliability and validity of the Addiction Severity Index with a homeless sample. J Subst Abuse Treat [Internet]. [cited 2019 Aug 23];11(6):541–8. Available from: http://www.ncbi.nlm.nih.gov/pubmed/7884837

41. Julious SA. Sample size of 12 per group rule of thumb for a pilot study. Pharm Stat. 2005;4(4):287–91.

42. National Center for PTSD. Using the PTSD Checklist for DSM-5 (PCL-5) [Internet]. [cited 2021 Sep 21]. Available from: https://www.ptsd.va.gov/professional/assessment/documents/using-PCL5.pdf

43. Barrett MS, Chua WJ, Crits-Christoph P, Gibbons MB, Thompson D. EARLY WITHDRAWAL FROM MENTAL HEALTH TREATMENT: IMPLICATIONS FOR PSYCHOTHERAPY PRACTICE. Psychotherapy. 2008.

44. Swift JK, Greenberg RP. Premature discontinuation in adult psychotherapy: A meta-analysis. J Consult Clin Psychol. 2012;

45. Voith LA, Hamler T, Francis MW, Lee H, Korsch-Williams A. Using a trauma-informed, socially just research framework with marginalized populations: Practices and barriers to implementation. Soc Work Res. 2020;

46. Bennett A, Crosse K, Ku M, Edgar N, Hodgson A, Hatcher S. Interventions to treat post-traumatic stress disorder (PTSD) in vulnerably housed populations and trauma-informed care: A scoping review. medRxiv. 2021;

